# Prevalence of HIV history and associated factors in people affected by mpox in Colombia, 2022

**DOI:** 10.1101/2025.02.05.25321718

**Authors:** Claudia Marcela Muñoz Lozada, Javier Alberto Madero Reales, Pablo Enrique Chaparro Narvaez, Franklyn Edwin Prieto Alvarado

**Author notes:** Corresponding author: (CM).

## Abstract

The 2022 mpox outbreak disproportionately affected men who have sex with men (MSM) and individuals with history of HIV. This study investigated the prevalence of HIV infection and associated factors among mpox cases in Colombia. A cross-sectional analysis of 4,046 confirmed mpox cases reported to the Public Health Surveillance System between June 25 and December 31, 2022, was conducted. The prevalence of a history of HIV infection in mpox cases was determined, and associated factors were assessed using bivariate and multivariate logistic regression. Most mpox patients were male (97%), with a median age of 31 years (IQR = 27, 37). The most affected age group was 25 to 34 years (52%), and 83.7% were MSM. The prevalence of HIV history was 60%, with the highest rates in Manizales, Medellín, and Itagüí. Common symptoms included genital lesions (59.6%) and fever (67.6%). Factors associated with HIV history in mpox patients were male sex [OR = 9.6, 95% CI = 4.35, 21.31], having sexually transmitted infections: hepatitis B [OR = 2.9, 95% CI = 1.91, 4.40], hepatitis C [OR = 7.0, 95% CI = 3.50, 13.97], syphilis [OR = 2.1, 95% CI = 1.46-3.12], hospitalization [OR = 2.2, 95% CI = 1.65, 2.93], Colombian nationality [OR = 2.5, 95% CI = 1.86, 3.48] and being homosexual [OR = 2.5, 95% CI = 2.09, 3.06]. A high prevalence of HIV infection (60%) was found among mpox patients in Colombia, predominantly among MSM. This highlights the need for targeted interventions, such as priority care for people with HIV, active surveillance, and education for timely diagnosis and prevention of complications.

## Introduction

Mpox is a disease caused by a viral agent belonging to the family *Poxviridae,* subfamily *Chordopoxvirinae* and genus *Orthopoxvirus* [1]. There are two lineages or clades of the virus, clade I (previously known as the Central African or Congo Basin clade) and clade II (previously known as the West African clade). The latter consists of two subclades IIa and IIb, to which the variants that circulated in the 2022 outbreak belong [2]. It is also estimated that clade I is more virulent than clade II [3].

Mpox is transmitted by direct contact with exudate from skin lesions, contact with feces of animals or sick people, inhalation of respiratory secretions, consumption of meat from wild animals or by contact with fabrics that have been in contact with a person or animal. sick *s* [1]. As of May 2022, mpox spread to more than 110 countries, reporting 84,100 cases, 67% in America, 32% in Europe and 1% in Africa; with characteristics and clinical manifestations different from the outbreaks described in previous years, related to mainly affecting young men, with transmission via direct physical contact including sexual contact, presentation of genital lesions and risk behaviors such as men who have sexual relations. sexual relations with other men (MSM) and a high presence of sexually transmitted diseases, identifying cases without a history of travel to endemic áreas [4–5] According to WHO data, 97% of cases occurred in men and 51 deaths were reported. In America, cases were reported in 24 countries; however, four countries accounted for 97% of the cases: the United States of America, Brazil, Canada and Peru. 82% of cases self-identified as MSM [6].

Changes in the epidemiology of the outbreak characterized by human-to-human transmisión were identified, which differs from traditional forms of transmission, active transmission in men and clinical characteristics such as the appearance of few skin lesions and frequent genital lesions, those that differ from the classic presentation [7]. Several reports agree that mpox occurs more frequently in men who have sex with men, who also present genital lesions, diagnosis of HIV and other sexually transmitted infections mpox [8–11]. According to the literature, for the evaluation of the clinical picture it is important to know the history of sexual contacts and history of contact with sick people, to make a differential diagnosis against pathologies related to sexually transmitted infections [5–12]. Due to the presentation of dermal lesions, it is necessary to perform a differential diagnosis with chickenpox and other diseases that present with skin lesions [9].

With the aim of understanding the sustained growth of mpox cases, Endo et al. (2022) carried out a statistical model using Weibull distributions adjusted to cases between 18 and 44 years of age in the United Kingdom, in this way, the dynamics of transmission through sexual contact networks were understood, identifying that a small number of cases in MSM is sufficient to cause an outbreak of large magnitude, given its basic reproduction number greater than 1 (R_0_ = 1.3) [13]. However, it is necessary to deepen the knowledge of the dynamics of propagation in networks of people, which limits the understanding of the particular phenomena of community transmission and in other groups of people, such as in the child population or pregnant women [14].

Several studies reported that spaces or events of mass influx have a close relationship with mpox infections, a situation that possibly explains the rapid spread worldwide. In a study in England, 64% of patients reported attending festivals, sex venues, private sex parties, or cruising venues (sexual activity with anonymous partners in public outdoor spaces) in the 21 days prior to the event. onset of symptoms [15]. Additionally, transmission can be sustained by contact with multiple partners, which makes it difficult to trace close contacts [15]. In another study, 58% of cases reported having visited clubs, bars or private parties during the infection period, which creates a challenge in establishing risk scenarios, due to multiple transmission networks [16]. In Spain, it was identified that the most common characteristics in 185 cases of mpox were drug use during sexual activity, HIV positivity and the diagnosis of another sexually transmitted disease [9].

In a study with reported cases of mpox in eight states of the United States in 2022, it was established that the prevalence of people with mpox who had an HIV infection was higher than the prevalence of HIV in the MSM population [11]. Also in the United States, Of 57 patients with severe manifestations of mpox, 82% (47/57) were positive for HIV, with almost a third receiving care at the ICU level and 21% of the patients dying [17]. Furthermore, in this same report, significant effects were evident on vulnerable populations, mostly Afro-descendant men and street dwellers who faced difficulties in accessing health services, health promotion and disease prevention activities, and to treatment for HIV [17–18].

With the increase in mpox cases in non-endemic countries, a problem arises that not only encompasses the health situation but also a social context that denotes a comprehensive approach to each individual, their family, partners, sexual contacts and social networks. [18]. This disease requires in-depth investigations into epidemiological, clinical and virological aspects since there are gaps in knowledge to determine the evolution of the disease and potential transmissibility [19]. Additionally, there is an urgent need to train health workers for public health response [19–20].

A challenge that takes form in containing mpox is the difficulties in early detection of cases, limiting chains of transmission and avoiding outbreaks in new cities [21–22]. On the other hand, there are challenges in the care of a disease with a high burden of stigma that affects the MSM community [22], which is why joint work between health personnel and patients is required to build an epistemology of self-care aimed at the MSM population [22]. The objective of this study was to determine the prevalence of HIV history and identify associated factors in those affected by mpox in Colombia between June 25^th^ and December 31^st^, 2022.

## Methods

### Study design and source of information

An analytical cross-sectional study was carried out, the information on mpox cases was obtained from the Colombian Public Health Surveillance System (Sivigila) to which laboratory confirmed cases are reported (RT-PCR). HIV history information was obtained from the historical HIV notification to Sivigila, which takes into account those with a laboratory-confirmed diagnosis, and in which their HIV status, AIDS stage, and AIDS death are reported only once. The High Cost Account (CAC) records the follow-up of the cohort of people living with HIV in Colombia, and the information related to the clinical stage and the treatment received by the patients. The study database was built from the information from the three mentioned sources.

### Population and sample

All people notified to Sivigila as mpox whose diagnosis was confirmed by laboratory test were included in the study (4,026). In addition, he was considered affected by mpox with a history of HIV to any person with a diagnosis of mpox confirmed by RT-PCR and with a history of HIV notified to Sivigila or registered in the CAC; and affected by mpox without a history of HIV to any person with a diagnosis of mpox confirmed by RT-PCR without a history of HIV notified to Sivigila or registered in the CAC. The records of those over 15 years of age, who met the case definition and who had a history of HIV through cross-checking with the Sivigila HIV or CAC databases were included. Records of affected people with incomplete information were excluded.

### Study variables

The information was organized in a Microsoft Excel spreadsheet including social and demographic variables (age, sex, social security affiliation, ethnicity, nationality, area of occurrence, socioeconomic stratum), epidemiological variables (travel history, close contact with person from abroad, close contact with a confirmed case in the last 21 days, new or multiple sexual partners, source of infection, sexual orientation and MSM) and clinics (hospitalization, final condition, date of onset of symptoms, date of skin rash, complications, HIV, other sexually transmitted infections) of those affected by mpox. To carry out the bivariate and multivariate analysis, the socioeconomic stratum was regrouped into low-middle (strata 1, 2 and 3) and high-middle (strata 4, 5 and 6), ethnic belonging into “others” and “black, mulatto, Afro-Colombian, Rom, gypsy and raizal” and age in the groups of 16 to 39 years and 40 and over.

### Statistic analysis

Prevalence was calculated with its respective 95% CI of the history of HIV in people affected by mpox, and the prevalence by departments and municipalities, as well as the monthly prevalence at the national level. Continuous quantitative variables with normal distribution were described using the mean and standard deviation. Quantitative variables that did not distribute normally were described in terms of median and interquartile ranges [23].

To explore the factors potentially associated with a history of HIV in those affected by mpox, a bivariate analysis was carried out between the dependent variable (history of HIV in those affected by mpox) and each of the independent variables (age, sex, MSM, hospitalization, STI, symptoms, mortality, stratum, nationality, area of occurrence, close contact with a person from abroad, social security, travel history, source of contagion, sexual orientation) using the Chi square test. Crude (cOR) and adjusted (aOR) odds ratios with their respective 95% CIs were calculated. The aORs were obtained through logistic regression analysis. All variables that were statistically significant in the bivariate analysis. To select the best model, the Akaike information criterion (AIC) was used to compare the fit of several models and identify the model that best fits the data (likelihood) and with the fewest number of parameters (complexity). Possible collinearity between the variables included in the final model was evaluated. Statistical significance was set at p < 0.05 Analyzes were performed using the R statistical language (R Core Team, 2023).

### Ethical considerations

The study was carried out in accordance with the Declaration of Helsinki and in accordance with resolution 8430 of 1993 and was classified as a risk-free study, since no intervention or treatment was performed on patients by the researchers. The interaction of the researchers was focused on the review of the databases as indicated in the methodology. For the study, information was used from a secondary source with anonymized data resulting from public health surveillance for Mpox and HIV. Anonymization involved the elimination of some variables such as names and identity data to ensure privacy and that the data does not allow the identification of individuals. Additionally, since the data is from an epidemiological surveillance system and within the framework of the public health emergency of international concern (PHEIC) due to the multi-country outbreak of Mpox in 2022, approval from the ethics committee was not required. The information obtained was used solely for public health purposes.

## Results

### Characteristics of the study population

4,046 individuals with laboratory-confirmed mpox were identified, of which 23 were younger than 16 years. Of the remaining 4,023 cases, 59.8% (2,408) had a history of HIV (Fig 1).

**Fig 1.**
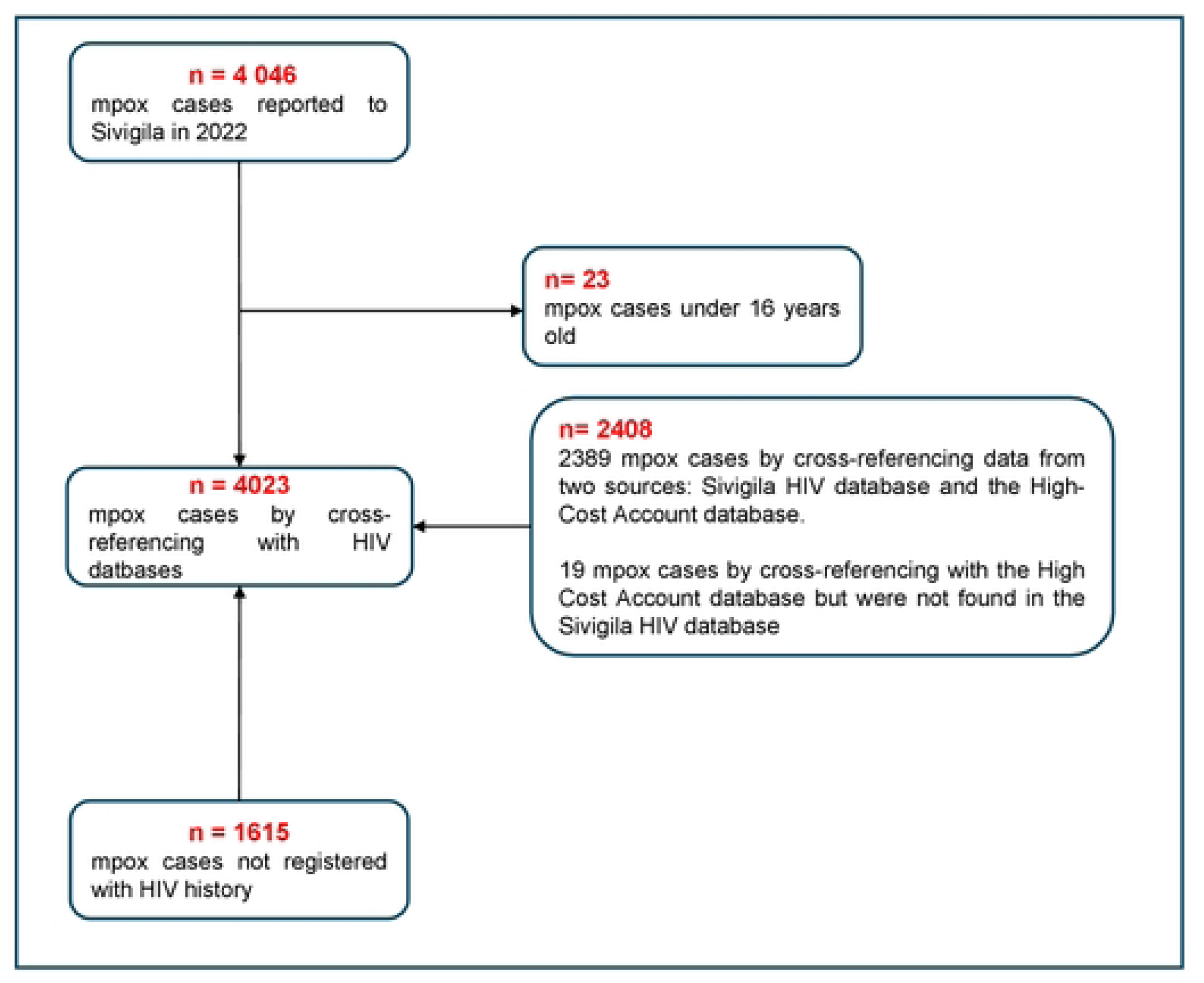
Flow diagram of the selection process for mpox cases with a history of HIV in Colombia, June to December 2022.

### Prevalence of HIV history in those affected by mpox in Colombia

The 4,023 cases considered were distributed in 123 municipalities that belong to 25 departments and 5 districts. The prevalence of HIV history in patients with mpox was 59.6% (2,410/4,046) (95% CI 58.0-61.1). During the beginning of the outbreak in Colombia, a considerable increase in the prevalence of the history of HIV in people affected by mpox, going from 20% to 71% between June and August 2023. Among the 15 municipalities that presented the highest number of cases, he highest prevalences were observed in the “coffee axis”: Manizales (72%) and Pereira (61.7%) and in the metropolitan area of Medellín (Medellín 71.8%, Itagüí 62.8% and Rionegro 60.9%). %). Municipalities with few reported cases had prevalences higher than 90% (Fig 2).

**Fig 2.**
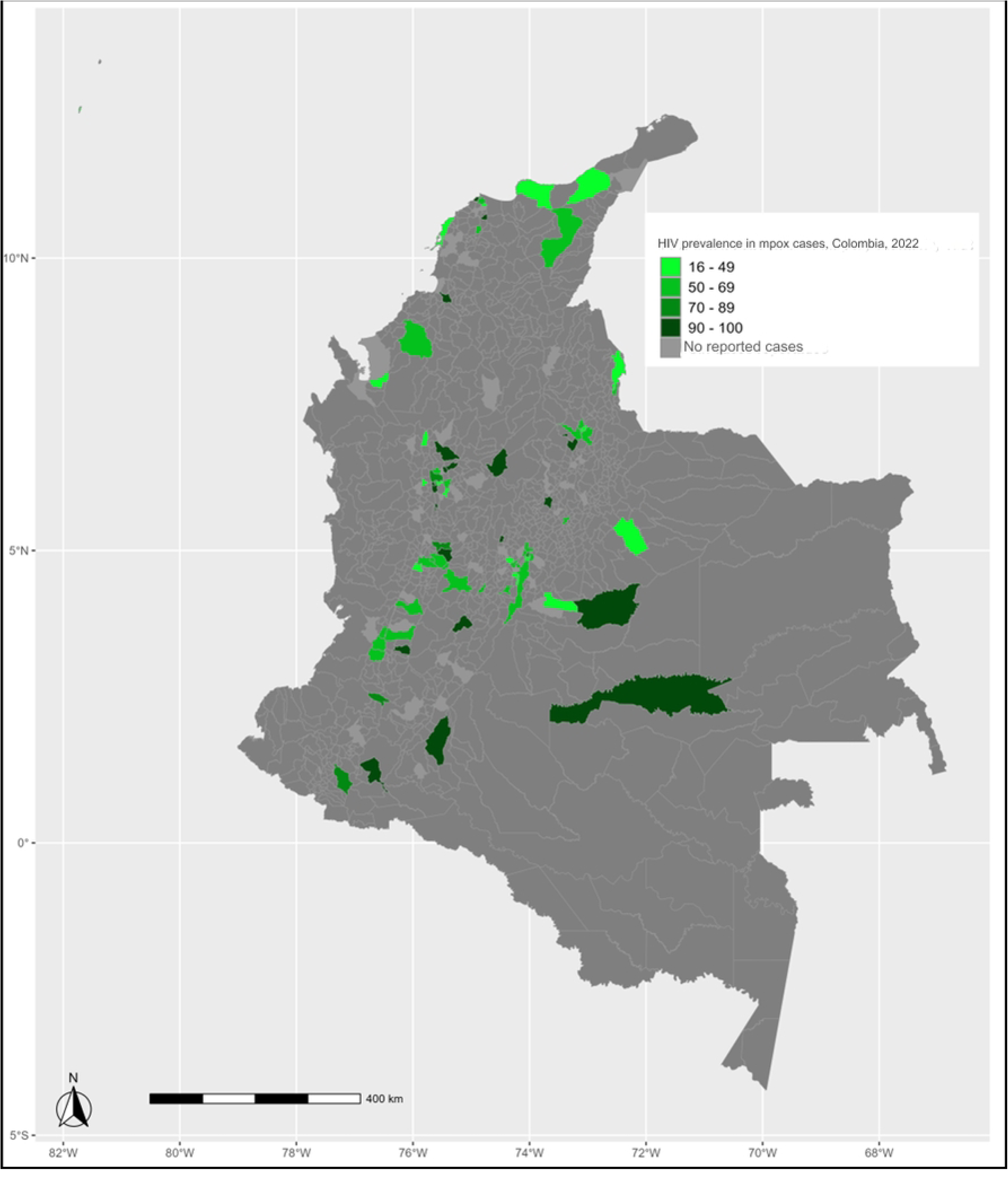
Prevalence of HIV history in those affected by mpox by municipality, Colombia, 2022.

Regarding those affected by mpox with a history of HIV, 2,408 (59.9%) were identified; of these, 99.5% (2,397) were men. By age groups, those most affected were those aged 25 to 29 years with 26.0% (625), 30 to 34 years old 28.6% (689) and 35 to 39 years old 18.6% (449).

The median age was 32 years (IQR 28-37). Regarding affiliation to the social security system, 79.4% (1,913) belonged to the contributory regime. Socioeconomic stratum 3 accounted for 39.3% (947). Regarding ethnicity, 99.8% (2,403) of those affected belonged to population groups that were not recognized as black, gypsy or raizal. 10.7% (257) of those affected were hospitalized and 0.4 (9) died (Table 1).

**Table 1.** Social and demographic characteristics of those affected by mpox with and without a history of HIV in Colombia, 2022.

| Variables | Confirmed cases mpox<br>(4,023) |  | Affected by mpox with a history<br>of HIV<br>(2,408) |  | Affected by mpox without HIV<br>history<br>(1,615) |  | p value |
| --- | --- | --- | --- | --- | --- | --- | --- |
|  | n | % | N | % | n | % |  |
| **Sex** |  |  |  |  |  |  |  |
| Male | 3,904 | 97.0 | 2,397 | 99.5 | 1,507 | 93.3 | **<0.001** |
| Female | 119 | 3.0 | eleven | 0.5 | 108 | 6.7 |  |
| **Age groups (years)** |  |  |  |  |  |  |  |
| 16-19 | 75 | 1.9 | 19 | 0.8 | 56 | 3.5 | **<0.001** |
| 20-24 | 513 | 12.8 | 214 | 8.9 | 299 | 18.5 |  |
| 25-29 | 1050 | 26.1 | 625 | 26 | 425 | 26.3 |  |
| 30-34 | 1043 | 25.9 | 689 | 28.6 | 354 | 21.9 |  |
| 35-39 | 650 | 16.2 | 449 | 18.6 | 201 | 12.4 |  |
| 40-44 | 349 | 8.7 | 218 | 9.1 | 131 | 8.1 |  |
| 45-49 | 176 | 4.4 | 110 | 4.6 | 66 | 4.1 |  |
| 50-54 | 108 | 2.7 | 62 | 2.6 | 46 | 2.8 |  |
| 55-59 | 3. 4 | 0.8 | 16 | 0.7 | 18 | 1.1 |  |
| 60 and over | 25 | 0.6 | 6 | 0.2 | 19 | 1.2 |  |
| **Affiliation with social security** |  |  |  |  |  |  |  |
| Contributory | 3,156 | 78.4 | 1913 | 79.4 | 1,243 | 77.0 | **<0.001** |
| Subsidized | 549 | 13.6 | 333 | 13.8 | 216 | 13.4 |  |
| Uninsured | 144 | 3.6 | 70 | 2.9 | 74 | 4.6 |  |
| Exception | 136 | 3.4 | 78 | 3.2 | 58 | 3.6 |  |
| Indeterminate | 19 | 0.5 | 7 | 0.3 | 12 | 0.7 |  |
| Special | 14 | 0.3 | 6 | 0.2 | 8 | 0.5 |  |
| Socioeconomic |  |  |  |  |  |  |  |
| 1 | 312 | 7.8 | 178 | 7.4 | 134 | 8.3 | 0.002 |
| 2 | 1,229 | 30.5 | 700 | 29.1 | 529 | 32.8 |  |
| 3 | 1,531 | 38.1 | 947 | 39.3 | 584 | 36.2 |  |
| 4 | 407 | 10.1 | 235 | 9.8 | 172 | 10.7 |  |
| 5 | 91 | 23 | 46 | 1.9 | Four. Five | 2.8 |  |
| 6 | 51 | 1.3 | twenty-one | 0.9 | 30 | 1.9 |  |
| Ethnicity |  |  |  |  |  |  |  |
| Others | 4,013 | 99.8 | 2,403 | 99.8 | 1,610 | 99.7 | 0.66 |
| Black, Afro-Colombian<br>mulatto | 7 | 0.2 | 4 | 0.2 | 3 | 0.2 |  |
| Rom, Gypsy | 2 | 0 | 1 | 0.0 | 1 | 0.1 |  |
| Raizal | 1 | 0 | 0 | 0 | 1 | 0.1 |  |
| Hospitalization |  |  |  |  |  |  |  |
| Yeah | 368 | 9.1 | 257 | 10.7 | 111 | 6.9 | <0.001 |
| No | 3,655 | 90.9 | 2,151 | 89.3 | 1,504 | 93.1 |  |
| Final condition |  |  |  |  |  |  |  |
| Alive | 4,012 | 99.7 | 2,399 | 99.6 | 1,613 | 99.9 | 0.137 |
| Deceased | eleven | 0.3 | 9 | 0.4 | 2 | 0.1 |  |
| Nationality |  |  |  |  |  |  |  |
| Colombian | 3,787 | 94.1 | 2,308 | 95.8 | 1,479 | 91.6 | <0.001 |
| Venezuelan | 223 | 5.5 | 95 | 3.9 | 128 | 7.9 |  |
| Other | 13 | 0.4 | 5 | 0.3 | 8 | 0.5 |  |
| Area of occurrence |  |  |  |  |  |  |  |
| Municipal head | 3,870 | 96.2 | 2,324 | 96.5 | 1,546 | 95.7 | 0.202 |
| Rural | 153 | 3.8 | 84 | 3.5 | 69 | 4.3 |  |
| **Population group** |  |  |  |  |  |  |  |
| Prisoner | 8 | 0.2 | 3 | 0.1 | 5 | 0.3 |  |
| Demobilized | 1 | 0.0 | 0 | 0.0 | 1 | 0.1 |  |
| Displaced | 1 | 0.0 | 1 | 0.0 | 0 | 0.0 |  |
| Disabled | 4 | 0.1 | 1 | 0.0 | 3 | 0.2 | <0.001 |
| Indigenous | 3 | 0.1 | 2 | 0.1 | 1 | 0.1 |  |
| Migrant | 161 | 4.1 | 64 | 2.7 | 97 | 6.1 |  |
| Victims of armed violence | 1 | 0.0 | 1 | 0.0 | 0 | 0.0 |  |
| Other population groups | 3,792 | 95.5 | 2,313 | 97.0 | 1,479 | 93.3 |  |

A 95.8 % (2,308) of those affected were Colombian nationals. Regarding the area of occurrence, 96.5% (2,324) of those affected appeared in the municipal seat. Regarding the population groups to which those affected belong, it was identified that 97% (2,313) belonged to the “others” population group.

### Bivariate analysis according to HIV history in patients with mpox

Compared to the comparison of proportions between those affected by mpox with a history of HIV and those affected by mpox without a history of HIV, there were significant differences in sex (p<0.001), age (p<0.001), affiliation to social security ( p<0.001), socioeconomic stratum (p <0.002), hospitalization (p<0.010), nationality (p <0.001) and population group (p <0.001) (Table 2).

**Table 2.** Clinical and epidemiological characteristics of those affected by mpox with and without a history of HIV in Colombia, 2022.

| Variables | Confirmed cases mpox<br>(4,023) |  | Affected by mpox with<br>a history of HIV<br>(2,408) |  | Affected by mpox<br>without HIV history<br>(1,605) |  | p value |
| --- | --- | --- | --- | --- | --- | --- | --- |
|  | N | % | N | % | n | % |  |
| Sexual orientation |  |  |  |  |  |  |  |
| Homosexual | 3,200 | 79.5 | 2,084 | 86.5 | 1,116 | 69.1 | <0.001 |
| Heterosexual | 577 | 14.3 | 244 | 10.1 | 333 | 20.6 |  |
| Bisexual | 166 | 4.1 | 78 | 3.2 | 88 | 5.4 |  |
| MSM |  |  |  |  |  |  |  |
| Yeah | 3,366 | 83.7 | 2,162 | 89.7 | 1,204 | 74.6 | <0.001 |
| No | 657 | 16.3 | 246 | 10.3 | 411 | 25.4 |  |
| Source of contagion |  |  |  |  |  |  |  |
| Unknown source | 3,246 | 80.7 | 1947 | 80.9 | 1,299 | 80.5 | 0.609 |
| Related to unknown source | 621 | 15.4 | 371 | 15.4 | 250 | 15.5 |  |
| Import related | 87 | 2.2 | 54 | 2.2 | 33 | 2.0 |  |
| Imported | 69 | 1.7 | 36 | 1.5 | 33 | 2.0 |  |
| Symptoms |  |  |  |  |  |  |  |
| Skin lesions on the body | 4,023 | 100.0 | 2,408 | 100 | 1,615 | 100.0 | 0.466 |
| Genital injuries | 2,400 | 59.7 | 1,460 | 60.6 | 940 | 58.2 |  |
| Fever | 2,722 | 67.7 | 1,626 | 67.5 | 1,096 | 67.9 |  |
| Asthenia | 1,217 | 30.3 | 718 | 29.8 | 499 | 30.9 |  |
| Headache | 1,094 | 27.2 | 606 | 25.2 | 488 | 30.2 |  |
| lymphadenopathy | 1,021 | 25.4 | 636 | 26.4 | 385 | 23.8 |  |
| Myalgia | 958 | 23.8 | 560 | 23.3 | 398 | 24.6 |  |
| Odynophagia | 794 | 19.7 | 461 | 19.1 | 333 | 20.6 |  |
| **New or multiple sexual partners** |  |  |  |  |  |  |  |
| Yeah | 480 | 11.9 | 272 | 11.3 | 208 | 12.9 | N/A |
| No | 671 | 16.7 | 339 | 14.1 | 332 | 20.6 |  |
| **Contact with mpox case in the last 21 days** |  |  |  |  |  |  |  |
| Yeah | 29 | 0.7 | 16 | 0.7 | 13 | 0.8 | 0.603 |
| No | 3,994 | 99.3 | 2,392 | 99.3 | 1,602 | 99.2 |  |
| **Travel history** |  |  |  |  |  |  |  |
| Yeah | 179 | 4.4 | 118 | 4.9 | 61 | 3.8 | 0.090 |
| No | 3,844 | 95.6 | 2,290 | 95.1 | 1,554 | 96.2 |  |
| **Close contact with a person from abroad** |  |  |  |  |  |  |  |
| Yeah | 516 | 12.8 | 306 | 12.7 | 210 | 13.0 | 0.783 |
| No | 3,507 | 87.2 | 2,102 | 87.3 | 1,405 | 87.0 |  |
| **Sexually transmitted infections** |  |  |  |  |  |  |  |
| Hepatitis C | 183 | 4.5 | 173 | 7.2 | 10 | 2.8 | <0.001 |
| Hepatitis B | 263 | 6.5 | 226 | 9.4 | 37 | 23 |  |
| Syphilis | 221 | 5.5 | 175 | 7.3 | 46 | 2.8 |  |

In individuals with a history of HIV affected by mpox, the main sexual orientation was being homosexual with 86.5% (2,084). 89.7% (2,162) of those affected were MSM. In 80.9% (1,947) the source of infection could not be identified. 1.5% (36) of those affected acquired the infection outside the country, of these, 25.0% (8) came from Spain and 21.9% (7) from the United States. Regarding symptoms, it was identified that all those affected had skin lesions, 60.6% (1,460) had lesions on the genitals, 67.5% (1,626) had fever and 29.8% (718) had asthenia. Complications were reported in three patients, two had a peritonsillar abscess and one had ophthalmic complications.

An 11.3% (272) reported a history of new or multiple sexual partners, 0.7% (16) reported contact with an mpox case in the last 21 days, 12.7% (306) reported close contact with person from abroad; 4.9% (118) reported a travel history. Regarding sexually transmitted infections, 7.2% (173) had hepatitis C, 9.4% (226) had hepatitis B, and 7.3% (175) had syphilis (Table 2). Compared to the comparison of proportions between the mpox group with a history of HIV and the mpox group without a history of HIV, the following variables were identified with significant differences: sexual orientation (p<0.01), MSM (p<0.01) and sexually transmitted infections (p<0.01) (Table 2).

Seven regression models were built and the selected one had an AIC of 3,939.1 and a deviation of 3,923 (Table 3). The final model explored seven factors that can explain the history of HIV in patients with mpox: being male (OR: 9.6; 95% CI: 4.35-21.31), hepatitis B (OR: 2.9; 95% CI:: 1.91-4.40), hepatitis C (OR: 7.0; 95% CI: 3.50-13.97), syphilis (OR: 2.1; 95% CI: 1.46-3.12), being hospitalized for mpox (OR: 2.2; 95% CI: 1.65-2.93), Colombian nationality (OR 2.5; 95% CI: 1.86-3.48) and being homosexual (OR: 2.5; 95% CI: 2.09-3.06)

**Table 3.** Multivariate model of the factors associated with mpox with a history of HIV, Colombia, 2022.

| Variables | OR | CI (95%) | p value |
| --- | --- | --- | --- |
| Sex |  |  |  |
| Man | 9.6 | 4.35-21.31 | < 0.001 |
| Women | Ref |  |  |
| Syphilis |  |  |  |
| Yeah | 2.1 | 1.46-3.12 | < 0.001 |
| No | Ref |  |  |
| Hepatitis B |  |  |  |
| Yeah | 2.9 | 1.91-4.40 | < 0.001 |
| No | Ref |  |  |
| Hepatitis C |  |  |  |
| Yeah | 7.0 | 3.50-13.97 | < 0.001 |
| No | Ref |  |  |
| Hospitalization |  |  |  |
| Yeah | 2.2 | 1.65-2.93 | < 0.001 |
| No | Ref |  |  |
| Homosexual |  |  |  |
| Yeah | 2.5 | 2.09-3.06 | < 0.001 |
| No | Ref |  |  |
| Colombian nationality |  |  |  |
| Yeah | 2.5 | 1.86-3.48 | < 0.001 |
| No | Ref |  |  |

OR: Odds Ratio, CI: Confidence Interval, Ref: reference

## Discussion

The present study found a prevalence of mpox patients with a history of HIV of 60%, and as associated risk factors a history of: hepatitis B, hepatitis C, syphilis; be hospitalized, Colombian nationality and have homosexual sexual orientation. In the mpox outbreak in 2022, Colombia ranked fifth in number of cases with 4,046 among the countries with the most reported cases, after the United States (30,611 cases), Brazil (10,967 cases), Spain (7,559 cases) and France (4,147 cases) [4]. Before 2022, several mpox outbreaks were described in Nigeria, the Democratic Republic of the Congo and the United States which affected few people [24–27].

In Colombia, the greatest impact occurred in homosexual men, this being similar to the findings of a study in New York, which reviewed records of the epidemiological surveillance of mpox and conducted interviews with 719 patients, identifying that 97.9% ( 704) were men, 78.7% (566) were gay, lesbian or queer; and 5.6% (40 cases) were bisexual, 77.6% (505) had intimate contact with men [28].

The prevalence of people affected by mpox with a history of HIV in Colombia was higher than those reported in other studies. In a study carried out in the states of California, Columbia, Georgia, Illinois and New York in the United States between May and July 2022, which included 1,969 affected people, the reported prevalence was 38% [29]. Another study in the United States, between May 17 and November 4, 2022, included 466 transgender and gender diverse adults, reporting a prevalence of 47.6% [30]. An investigation in Brazil, in a hospital in Rio de Janeiro between June and August 2022 with 205 cases indicated 53% [31], while another in Nigeria carried out between 2017 and 2018 that included 40 cases calculated a prevalence of 22.5% [27]. On the other hand, a systematic review that included 53 studies with confirmed cases from 16 countries published since the beginning of the outbreak in multiple countries and until October 1, 2022, reported 6,345 individuals and reported a prevalence of 40.3% [32].

The differences noted between the prevalence found in Colombia, with respect to the aforementioned studies, may be related to the availability of the two sources with historical information and with national scope that were consulted to verify the history of HIV infection; Additionally, this study included information from mpox case notifications. Studies had already indicated that these infections are cofactors to facilitate the transmission of mpox or HIV infection, taking into account that the immunosuppression caused by HIV generates important changes in the clinical course of mpox disease, which manifests in a longer illness and greater number and size of skin lesions and more frequent appearance of genital lesions [27].

In the population notified with HIV in Colombia The impact by mpox was disproportionate. This population in 2022 was 141,787 people, and is made up of a large proportion of MSM (44% (62,386) in people living with HIV by 2022 in Colombia), and who have higher risk sexual behaviors such as having multiple partners and in many cases risky sexual contacts with unknown partners [33].

The findings of mpox with a history of HIV by age groups in this study show greater involvement in those aged 25 to 29 years (26.0%), and those aged 30 to 34 years (25.8%), while in another study in Brazil, the most affected were men between 30 and 39 years old [31], which is the economically active population; Possibly these age groups actively participate in networks of sexual contacts that facilitate and promote the rapid transmission of mpox infection, especially in spaces of casual sexual encounter, similar to what was found in an ethnographic study in Argentina, which describes the practices sexual relations and meeting places [34].

Homosexual and bisexual men and those in the key group of MSM are the most affected by mpox infection, possibly because they could be in sexual networks where the highest incidence of mpox occurs [30]. In this study, the prevalence of a history of HIV in those affected by mpox in the key group of MSM is 64%, being higher than the prevalence of HIV in the group of MSM (44.2%) when taking as reference the number of people living with HIV in Colombia in the Colombia High Cost Account report [33].

In Colombia, the high prevalence of HIV history in MSM with mpox not only indicates that this population, possibly due to their practices of greater intimate or sexual contact, faces a greater risk of contracting mpox infection, but also that they face the stigma and discrimination that they have historically received due to their status as a person living with HIV. This key group has been blamed for the spread of the global HIV epidemic, and now, due to the stigma generated by the concentration of mpox cases in this key group, visits to health services may decrease and the effectiveness of measures, such as vaccination, early care of HIV patients with complications, and educational campaigns may be affected [35].

In this study, no significant differences were found in the presence of symptoms in mpox patients with a history of HIV, possibly because the HIV cases were receiving antiretroviral therapy, which could have contributed to the mild symptoms observed. This situation differs with several studies, one of them carried out in 2023 that used as a source of information the registry of mpox patients from 29 countries that were registered in the surveillance system [1] and which reported that HIV patients confirmed for mpox had greater probabilities of having diarrhea (p=0.002), skin rash or perianal lesions (p=0.03) and a greater rash burden (p < 0.001) [1].

In the present study, it was identified that 10.7% (257) of patients with mpox with a history of HIV were hospitalized and 0.4% (9) died. This finding differs from that of a study carried out in patients with mpox and HIV. in 19 countries that reported that 28% (107) were hospitalized and of them 27 (25%) died. More deaths occurred in people with CD4 counts less than 200 cells per mm³ [36]. Likewise, another study showed that people with HIV infection and mpox have a higher proportion of hospitalization (8% versus 3%) [29].

The cities with the highest prevalence of mpox infection with a history of HIV (higher than the national prevalence) were Manizales, Medellín, Itagüí, Pereira and Rionegro, a situation that can be explained because the largest number of people living with HIV are registered in the central region of Colombia prevails [33], where these cities are located; allowing the network of these people to present greater interactions or contacts that help the transmission of mpox.

The population with HIV presents inequities in accessing the health system, in terms of specialized consultations, not to mention access to pre-exposure prophylaxis (PrEP) to HIV, which, in the case of Colombia, is still associated with high costs and not It is still included in individual health benefit plans. However, the above, in the sources of the study, the use of PrEP by people with mpox was not systematically, reliably and completely included, which becomes relevant when, within these sexual networks, the use of these medications has It has allowed people living with HIV to improve their quality of life, especially in high-income countries, where the debate on risk compensation is revived, taking into account, for example, the decrease in the frequency of condom use in the MSM population [37].

The associated factors explored in individuals affected by mpox with a history of HIV were found to be risky: *being a man, having STIs such as syphilis, hepatitis B, hepatitis C, being hospitalized, Colombian nationality and being homosexual. These factors are consistent with those reported by a literature review carried out in Europe where being a young man, having sex with other men, HIV positivity and a history of* STIs [5–12] were identified as preliminary risk factors. HIV can be configured as a marker of higher risk behavior that facilitates the acquisition of mpox, by globally identifying a greater proportion of men with confirmed mpox [8].

These similarities between the associated risk factors found in Colombia and in other studies suggest a strong relationship between the sources of contagion and the complex networks that are identified in the HIV and MSM population. mm³ [36]. Networks of casual sexual encounters and contacts in the community of people living with HIV and in homosexual people constitute an opportunity for the agent (mpox) to be transmitted as interactions and intimate contact increase. human that allows the impact of a greater number of people who are part of that network.

However, the same complexity and structure of the network limits the capacity to affect other groups in the community that do not participate in this network. The network of people living with HIV behaves as a complex network, and in the context of the transmission of this disease, social and cultural factors influence such as participation in networks for sexual encounters with unknown people and without the use of a condom. These factors are not static over time, the transmission of HIV is not only due to its effectiveness, nor to its latency period, but above all to the complex structure of this network due to its sexual practices [38].

In order to understand more broadly a complex network, such as the one present in people living with HIV, it is necessary to make use of complementary information either from secondary sources, such as the High Cost Account, or better from primary sources where Through a qualitative research approach, the feelings and presence of those affected can be understood, who, as can be seen in the study and given the high prevalence of pre-existing HIV infection, are individuals who still suffer from stigma. and discrimination due to their condition.

The study identifies limitations related to the use of secondary sources; it is possible that the prevalence found would be higher if the care protocol for mpox cases required laboratory HIV testing at the time of the consultation. It is possible that in this study there was under-reporting in the notification of patients affected with mpox with mild symptoms, who decided not to consult health services; On the other hand, the data sources where the history of HIV was identified (Sivigila and CAC) may also have underreported or not have the most recent information on the history of HIV; It is possible that some mpox-affected patients were HIV positive but were not identified in this study.

To reduce potential selection bias, inclusion criteria were strictly applied. Regarding classification bias, the bases for performing classification according to the established case definition were reviewed. Confounding factors were avoided by reviewing laboratory diagnosis dates to select positive cases. A possible confounding factor in this research could be given by the age of the patients, since the young adult population has a greater propensity to engage in risk behaviors related to greater exposure to sexually transmitted diseases, unlike, for example, the population of older adults [39].

It was identified that the highest prevalence corresponded to the municipalities of Manizales, Medellín, Itagüí, Pereira and Rionegro. The most affected were men between 25 and 39 years old, the most frequent symptoms and signs were skin lesions, genital lesions, fever and asthenia. For those affected by mpox with a history of HIV, being a man, having sexually transmitted infections such as hepatitis B, hepatitis C and syphilis, being hospitalized, Colombian nationality and being homosexual were identified as risk factors.

For those affected by mpox with a history of HIV, the Colombian health system must ensure the availability and access to diagnostic tests, prevention and health education strategies, and timely treatments as a priority. Health service providers and insurers must perform tests for HIV diagnosis along with CD4 cell count and tests for the identification of other STIs when there is suspicion and confirmation of mpox cases in the country.

The main population in Colombia affected by mpox were young men who have relationships with other men and a high proportion has a history of HIV, a situation that establishes the need to carry out communication and education strategies at the territorial level, specifically aimed at this population and that Allow the participation of leaders and the general population.

An approach is required from the perspective of public health that identifies the needs of the population at high risk, through participatory scenarios that allow the promotion of public health actions at all levels and the development of other research that expands information on complications of patients over time, as well as going deeper with qualitative studies that allow us to understand the dynamics of the health and disease process, for a better understanding and approach to this disease.

## Data Availability

All relevant data are within the manuscript and its Supporting Information files.

## Acknowledgment

To all the surveillance teams at the departmental and district level that led the response to the mpox outbreak in Colombia and to the Virology Laboratory Group of the National Institute of Health.

## Financing

This study was funded by the Directorate of Surveillance and Risk Analysis in Public Health of the National Institute of Health and the Pontificia Universidad Javeriana.

